# Associations of Anxiety, Depression, and Sleep Quality with MNBI, PSPW-I, and ECJ-CI in rGERD, FH, and RH Patients: A Retrospective Study

**DOI:** 10.1101/2025.02.20.25322595

**Authors:** Lunsheng Lou, Yurong Huang, Wangliu Yang, Gengqing Song, Jie Yang

**Author notes:** These authors contributed equally to this work (Lunsheng Lou, Yurong Huang).

## Abstract

**Objective:** This study examined the impact of anxiety, depression, and sleep quality on mean nocturnal baseline impedance (MNBI), post-reflux swallow-induced peristaltic wave index (PSPW-I), and esophagogastric junction contractile index (EGJ-CI) in patients with refractory gastro-esophageal reflux disease (rGERD), functional heartburn (FH), and reflux hypersensitivity (RH).

**Methods:** Retrospective analysis included 75 patients aged 18-70 with persistent reflux symptoms despite 8 weeks of proton pump inhibitor (PPI) therapy. Evaluations included esophagogastroduodenoscopy, high-resolution manometry, 24-hour pH-impedance monitoring, and psychological assessments (SAS, SDS, PSQI). Patients were grouped into rGERD, FH, and RH. MNBI, PSPW-I, and EGJ-CI were compared, and correlations with psychological and sleep parameters were analyzed.

**Results:** Distal MNBI was significantly lower in rGERD (1177.91 ± 707.22 Ω) vs. FH (1995.77 ± 476.02 Ω) and RH (2062.35 ± 509.93 Ω) (P < 0.001). Anxiety prevalence was 85.7% in rGERD, 64.7% in FH, and 67.5% in RH (P = 0.302); depression affected 78.6% of rGERD, 70.8% of FH, and 72.9% of RH patients (P = 0.942). Poor sleep quality was present in >80% of all groups. PSPW-I negatively correlated with anxiety (r = −0.181, P < 0.05) and depression (r = −0.158, P < 0.05), as did proximal MNBI with depression (r = −0.175, P < 0.05). A distal MNBI cutoff of 1531 Ω distinguished rGERD from FH/RH with 71.4% sensitivity and 87.5% specificity (AUC = 0.80).

**Conclusion:** Anxiety, depression, and poor sleep quality impair PPI efficacy and worsen esophageal acid clearance. Distal MNBI effectively differentiates rGERD from FH/RH. Addressing psychological and sleep disturbances may improve treatment outcomes in refractory reflux patients.

## Introduction

Inadequate resolution of reflux symptoms with proton pump inhibitors (PPIs) necessitates consideration of other conditions such as refractory gastro-esophageal reflux disease (rGERD), functional heartburn (FH), and reflux hypersensitivity (RH). Multichannel intraluminal impedance combined with pH monitoring (MII-PH) parameters such as mean nocturnal baseline impedance (MNBI) and post-reflux swallow-induced peristaltic wave index (PSPW-I) are reflective of esophageal mucosal integrity and acid clearance[1]. Esophagogastric junction contractile index (EGJ-CI) that quantifies gastroesophageal junction function is a novel high-resolution esophageal manometry (HREM) parameter that has been proved valuable in distinguishing between rGERD, FH, and RH[2] Accumulating evidence indicates that anxiety, depression, and sleep disorders can induce esophageal histopathological changes and esophageal motility disorders and reduce the effectiveness of PPI therapy in reflux patients with reflux symptoms[3]. However, their influence on parameters such as MNBI, PSPW-I, and EGJ-CI is yet unclear.

This study analyzed MNBI, PSPW-I, and EGJ-CI among patients with rGERD, RH, and FH, and investigated the impact of anxiety, depression, and sleep quality on these parameters. The aim was to improve the existing knowledge of these new indicators in the management of reflux-like symptoms.

## Subjects and Methods

### Study subjects

Clinical data were collected for patients who presented with the following symptoms at the Liupanshui People’s Hospital from September 2021 to June 2023 and then retrospectively analyzed: acid regurgitation, heartburn, chest pain (discomfort behind the sternum).the authors did not have access to any information that could identify individual participants during or after data collection.

### Inclusion criteria

Patients were included in the study if they met the following criteria: symptoms of heartburn, acid regurgitation, chest pain or discomfort behind the sternum (non-cardiac chest pain) that failed to improve or only partially improved after standard-dose PPI therapy for 8 weeks[4, 5]; age between 18 and 70 years.

### Exclusion criteria

Patients were excluded based on the following criteria: those allergic to PPIs; patients unwilling to undergo gastroscopy and esophageal high-resolution manometry with impedance-pH monitoring; patients with a history of gastric or esophageal tumors or surgery; patients diagnosed with eosinophilic esophagitis; those with severe esophageal motility disorders.

### Investigations

All Prior to study participation, all patients discontinued antisecretory medications and prokinetic agents for at least 1 week prior to study participation. Each patientand underwent the following evaluations:

1. Gastroduodenoscopy
2. Esophageal high-resolution manometry (gastroduodenoscopy, HREM)
3. 24-hour impedance-pH monitoring.
4. Assessment tools
5. Patients completed the following self-rating scales:
6. Zung Self-Rating Anxiety Scale (SAS); Zung Self-Rating Depression Scale (SDS); Pittsburgh Sleep Quality Index (PSQI).

### Diagnostic criteria

Patients were categorized into three groups based on the following criteria: rGERD, acid exposure time (AET) > 6.0% or 4.0% ≤ AET ≤ 6.0% (abnormal total reflux events or positive reflux-symptom association); FH, AET < 4.0%, normal total reflux events, and negative reflux-symptom association; RH, AET < 4.0%, normal total reflux events, and positive reflux-symptom association[6].

### Liquid-state HREM

Patients were instructed to fast for at least 8 hours prior to examination. While in seated position, a lubricated catheter was inserted through the patient’s nostril into the esophagus. After a 3-5 minute adaptation period, data collection was commenced. Patients were instructed to breathe calmly and refrain from swallowing for 30 seconds. Resting pressure of the lower esophageal sphincter (LES) was recorded, and then the patient was asked to swallow 5 mL of warm water in supine position for 10 times[7].

A distal contractile index (DCI) tool was used to calculate values of esophagogastric junction (ECJ). The values that were 2 mmHg higher than the intragastric pressure over three consecutive respiratory cycles were divided by the duration (s) of three consecutive respiratory cycles and expressed in mmHg·cm[6]. Pressure topography maps of the esophagus were automatically generated using a pressure measurement system, and then analyzed as per the Chicago Classification 4.0 for esophageal motility disorders.

HREM parameters included upper esophageal sphincter pressure, lower esophageal sphincter pressure, distal contraction integral, and distal latency.

### 24-hour impedance-pH monitoring

Following HREM, portable multi-channel impedance-pH monitoring (China, Jinshan) was conducted. In brief, pH sensors were positioned 5 cm above the LES and six impedance probes were placed at 17, 15, 9, 7, 5, and 3 cm above the LES[7]. The patients maintained their usual activities over 24 hours. Data recorded included meal time, sleep (supine position), wake up time, symptom onset time (e.g., heartburn, acid regurgitation, chest pain or discomfort behind the sternum, belching, coughing, etc.), and medication intake time. The automated software of the monitoring system calculated AET, DeMeester score, number and types of reflux events, prolonged reflux events, and duration of the longest reflux event. From impedance-pH monitoring curves, three time points (1:00 AM, 2:00 AM, 3:00 AM) were selected to assess MNBI (average values were considered to avoid pH value fluctuations caused by swallowing or reflux). Proximal MNBI was measured as the average of the proximal two channels, while distal MNBI was calculated as the average of the distal four channels[8].

PSPW during MII-pH monitoring was defined as a swallow event occurring within 30 seconds after restoration of impedance to baseline levels at the most distal channel (complete bolus clearance) in response to a reflux event (decrease in sequential impedance values of 50% from the most proximal to the most distal channel)[9]. PSPW-I was derived by dividing the number of PSPWs by the number of reflux events.

Subjects were categorized into rGERD, FH, and RH groups based on AET, number of reflux events, and their correlation with symptoms. AET indicates the percentage of time with distal esophageal pH < 4 over 24 hours and was interpreted as follows: AET > 6.0%, pathological acid exposure; < 4.0%, normal; 4.0%-6.0%, uncertain. Reflux events were classified as acidic (pH < 4), weakly acidic (pH 4-7), and alkaline (pH > 7). Overall, >80 reflux events were considered abnormal[6].

Symptom association probability (SAP) and symptom index (SI) were used to assess the correlation between symptoms and reflux events. An SI ≥ 50% and/or SAP ≥ 95% indicated a positive reflux-symptom association.

### Anxiety, depression, and sleep quality assessment

Anxiety and depression were assessed using SAS and SDS, respectively. Each scale comprised 20 questions. Anxiety scores were evaluated as follows: score < 50, normal; 50-60, mild anxiety; 61-70, moderate anxiety; 70 or more, severe anxiety[10]. Depression scores were evaluated as follows: score < 53, normal; 53-62, mild depression; 63-72, moderate depression; 72 or more, severe depression[11]. PSQI was used to evaluate sleep quality across seven dimensions, namely, subjective sleep quality, sleep latency, sleep duration, habitual sleep efficiency, sleep disturbances, use of sleeping medication, and daytime dysfunction. Scores ranged from 0 to 21, with higher scores indicating poorer sleep quality[12]. All patients completed the aforementioned assessments.

### Statistical analysis

All statistical analyses were performed using SPSS 26.0. Normally distributed continuous data were expressed as mean ± standard deviation (x ± s). One-way analysis of variance (ANOVA) was used for comparisons among three groups, which was followed by pairwise comparison if P < 0.05. Continuous data with skewed distribution were expressed as median (interquartile range), and comparisons between three groups were performed using Kruskal-Wallis H test, which was followed by Bonferroni correction for post hoc comparisons if P < 0.05. Categorical data were presented as counts (percentages), and group comparisons were made using a chi-square test or Fisher’s exact test. Receive operating characteristic (ROC) curve analysis was performed to evaluate the diagnostic value of distal MNBI in distinguishing rGERD from FH and RH. Spearman analysis was employed to explore correlations between HREM parameters, 24-hour impedance-pH monitoring parameters, and anxiety/depression. Statistical significance was set at P < 0.05 for all analyses.

## Results

### Clinical characteristics of the patients

A total of 75 patients were enrolled, including 36 male and 39 female patients between 18 and 65 years of age. Of these, 14 patients had rGERD, 24 had FH, and 37 had RH. No statistically significant differences were observed in age, gender, duration of illness, heartburn, and chest pain (retrosternal discomfort) between the three groups. The body mass index (BMI) was significantly higher (P < 0.001) for patients with rGERD than for those with FH and RH (overweight was defined as a BMI value of 25-29 kg/m²). Acid reflux was significantly more common in rGERD patients (P < 0.05) than in the other two groups (S1 Table).

### HREM and MII-PH monitoring

The value of EGJ-CI reported in rGERD patients was lower than that reported for FH and RH patients; no difference was observed in EGJ-CI between FH and RH patients. Proximal MNBI showed no significant difference between the three groups; however, distal MNBI was significantly lower in rGERD patients than in FH and RH patients. There was no statistically significant difference in PSPW-I among patients with rGERD, FH, and RH (S2 Table).

### Anxiety, depression, and sleep quality

Anxiety and depression are very common among patients with refractory reflux symptoms, typically observed in more than 60% of patients. Anxiety was more common than depression in patients with rGERD, FH, and RH; however, the percentages of patients with anxiety and depression were not statistically different between the three groups. A total of 85.7%, 64.7%, 67.5% patients with rGERD, FH, and RH, respectively, had anxiety (P = 0.302^α^), while 78.6%, 70.8%, and 72.9% patients reported depression (P = 0.942^α^). Sleep quality was good in less than 20% of patients from the three groups (14.3%, 20.8%, 18.9% for rGERD, FH, and RH, respectively; P = 1.000^α^). ‘Poor’ sleep quality was more common than anxiety and depression in the three groups (Table 3).

### Correlation analysis

Spearman’s correlation analysis revealed the negative correlation between PSPW- I and anxiety and depression (r = −0.181 and −0.158, respectively; both P < 0.05). Further, proximal MNBI was found to be negatively correlated with depression (r = −0.175, P < 0.05) (S4 Table).In the ROC analysis, the area under the curve of distal MNBI was 0.80, the cutoff value was 1531 Ω, the sensitivity was 71.4%, and the specificity was 87.5%. Distal MNBI could significantly differentiate patients with rGERD from patients with HF and RH (Fig 1).

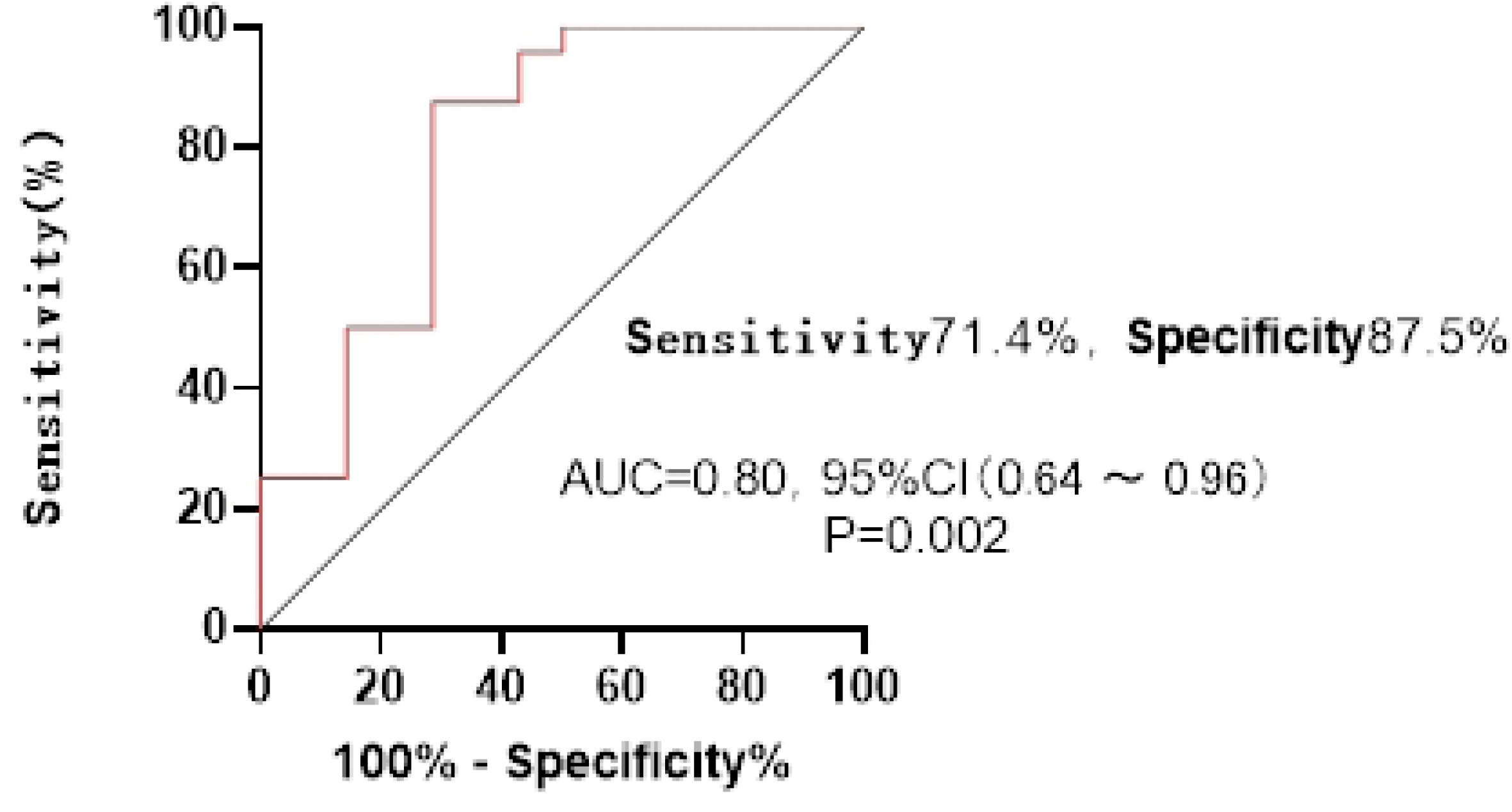

## Discussion

The reflux-like symptoms of gastroesophageal reflux disease (GERD) are complex and varied, including esophageal symptoms, such as acid regurgitation, heartburn, chest pain, and dysphagia, as well as extra-esophageal symptoms, such as cough, hoarseness, asthma, and dental erosion. Proton pump inhibitors (PPIs) are the first-line treatment for GERD; however, in patients who do not respond effectively, a condition often referred to as refractory GERD, both domestic and international guidelines recommend further clarification to determine whether the patient truly has GERD. These patients may be ultimately diagnosed with true GERD, functional heartburn, reflux hypersensitivity, gastric belching, or rumination syndrome[3].

The Lyon Consensus of 2018[13] defined functional heartburn (FH) as normal acid exposure, with no pathological reflux, and a negative correlation between reflux and symptoms, whereas reflux hypersensitivity (RH) was defined as normal acid exposure, no pathological reflux, and a positive correlation between reflux and symptoms.

In this study of 75 patients, 33% were diagnosed with FH, 48% with RH, and only 19% with true GERD. Among the true GERD cases, 71.4% were classified as NERD, indicating that NERD is more common than RE in rGERD, consistent with the findings of many studies[4, 14]. Furthermore, this study further confirmed that non-acid reflux is more common in patients with rGERD-like symptoms. A 24-hour pH-impedance test can assist in the prompt adjustment of treatment strategies, rather than continuing to intensify acid suppression therapy.

In the present study, more than 90% of patients with rGERD exhibited symptoms of acid regurgitation, in contrast to the FH and RH groups where the frequency of acid regurgitation was lower. This suggests that the absence of major acid regurgitation symptoms may indicate a lower likelihood of true GERD. Therefore, in clinical practice, healthcare institutions that lack reflux monitoring facilities can use the presence or absence of acid regurgitation as a simple method for the identification of true GERD, potentially improving treatment outcomes through optimized acid suppression strategies.

The difficulty in relieving reflux symptoms may be influenced by several factors: impaired esophageal mucosal integrity, decreased esophageal chemical clearance, dysfunction of the gastroesophageal junction (GEJ) barrier, comorbid mental health disorders, esophageal hypersensitivity, and brain-gut axis dysregulation[3].

In patients with NERD, normal endoscopic findings suggest that the primary cause of heartburn is a functional defect in the GEJ barrier, which cannot be detected endoscopically[4]. This includes the presence of dysfunctional tight junctions between cells, reduced mucosal protective factors (e.g., insufficient mucus and bicarbonate secretion), and impaired cell regeneration and repair. These changes are not sufficient to cause visible endoscopic lesions, despite potentially leading to changes in the mucosal impedance (MNBI). In our study, patients with true GERD showed significantly lower distal MNBI values compared to those with FH and RH, indicating compromise of the esophageal mucosal barrier in true GERD, while the integrity of the mucosa is relatively intact in patients with FH and RH. This suggests that symptoms associated with FH and RH are more likely to originate from esophageal hypersensitivity, brain-gut axis dysfunction, and other factors[15] There were no significant changes in the proximal MNBI values among the true GERD, RH, and FH groups in the present study, suggesting that this marker is not useful in distinguishing between the three. Frazzoni et al.[16] found that a baseline impedance value of less than 1500 Ω supports the diagnosis of GERD. The Lyon Consensus 2.0[6] proposed that MNBI values below 1500 Ω at 3 and 5 cm above the lower esophageal sphincter (LES) indicate impaired esophageal mucosal integrity. In our results, the optimal cutoff for distal MNBI in distinguishing rGERD from FH and RH was 1531 Ω, with a sensitivity of 71.4% and specificity of 87.5%. This further confirms the importance of distal MNBI in the differentiation of rGERD and warrants further verification in multi-center, large-scale studies.

In GERD and/or rGERD, a reduction in the post-deglutitive swallow-induced pressure wave index (PSPW-I) indicates reduced esophageal chemical clearance[17]. Ribolsi et al.[9] showed that a decrease in PSPW-I in rGERD results in a prolonged contact of acid, weak acid reflux, or even physiological reflux, stimulating the secretion of pro-inflammatory cytokines by the esophageal epithelium, inducing epithelial proliferation, and recruiting T lymphocytes and other inflammatory cells, ultimately resulting in mucosal damage. Furthermore, decreased PSPW-I leads to activation of sensory nerve endings expressing acid-sensitive ion channels, resulting in reflux symptoms[18].

In our results, the PSPW-I values in patients with rGERD, FH, and RH were 6.2%, 6.1%, and 5.5%, respectively, significantly lower than the 61% observed in healthy subjects[17]. This suggests that the reduction in PSPW-I is a key contributors to rGERD symptoms, consistent with the findings of Frazzoni et al. [17]. However, PSPW-I is unable to distinguish effectively between true GERD, FH, and RH.

Impaired barrier function at the gastroesophageal junction (GEJ) leads to failure in preventing the entry of gastric and duodenal contents into the esophagus and is one of the primary causes of GERD. Wang et al.[19] found that the EGJ contractile index (EGJ-CI) measured by high-resolution esophageal manometry could effectively differentiate rGERD from FH using a cutoff value of 35.8 mmHg·cm. Our study further demonstrated that patients with rGERD had significantly lower EGJ-CIs than patients with FH or RH, while no significant difference was observed between FH and RH. This suggests that the barrier dysfunction at the GEJ is more pronounced in true GERD than in FH and RH. psychological comorbidities, particularly anxiety and depression, indicating potential reciprocal causation[20].The study revealed distinct anxiety prevalence rates across the study groups: 85.7% in refractory GERD patients, compared to 64.7% in functional heartburn (FH) and 67.5% in reflux hypersensitivity (RH) patients. In contrast, depression rates showed comparable patterns among groups, with 78.6% in rGERD, 70.8% in FH, and 72.9% in RH patients, without reaching statistical significance.In accordance with previous research findings, our study corroborates the high prevalence of psychological disturbances, specifically anxiety and depression disorders, in individuals with refractory GERD manifestations.Comparative analysis demonstrated comparable clinical characteristics across different phenotypic subgroups, without reaching statistical significance[21].The study revealed that psychological determinants potentially outweigh physiological indicators in their impact on symptom severity assessment[15].The mechanisms responsible for the limited effectiveness of proton pump inhibitor therapy are multifactorial and complex.

Ha et al.[22] found that effective treatment of GERD is essential for reducing comorbidities related to poor sleep quality. In our study, impaired sleep quality was common in rGERD patients, with 85.7%, 79.1%, and 81.0% of patients in the three groups, respectively, reporting “poor” sleep quality. However, the severity of sleep disturbances did not differ significantly between the groups. This is consistent with the findings of Geeraerts et al.[18] and suggests that treatments aimed at improving sleep quality may benefit overall rGERD symptoms.

Although anxiety, depression, and poor sleep quality are commonly observed in patients with rGERD, their effects on the new esophageal diagnostic indicators are not yet well understood. Huang et al. [23] found that treatment of comorbid anxiety and depression improved the outcomes of rGERD therapy. This study identified a weak but statistically significant inverse correlation between depression levels and proximal mean nocturnal baseline impedance (MNBI), potentially indicating an association between depressive states and compromised proximal esophageal mucosal integrity.The analysis revealed weak but statistically significant negative correlations between both anxiety and depression scores with post-reflux swallow-induced peristaltic wave index (PSPW-I), suggesting that psychological factors may potentially impair esophageal chemical clearance efficiency.Our findings suggest that adjunctive treatment of anxiety and depression disorders may contribute to improved therapeutic responses in rGERD, potentially through dual mechanisms involving the reinforcement of esophageal anti-reflux barrier integrity and optimization of chemical clearance capacity.

The present study found that poor sleep quality was not significantly associated with proximal MNBI, distal MNBI, EGJ-CI, or PSPW-I, suggesting that improving the esophageal barrier and clearance functions may not directly affect sleep quality. Sleep quality is influenced by many factors beyond esophageal function, such as psychological state, weight, age, and lifestyle habits[24]. In other words, a simple improvement in sleep quality may not be sufficient improve esophageal function. Given that sleep disturbances are common in rGERD, they may increase esophageal acid exposure via centrally mediated esophageal hypersensitivity and the effects of satiety hormones[24]. The correction of sleep disturbances may therefore contribute to improving overall GERD symptoms.

This study has the following limitations: the study did not include healthy volunteers, making it difficult to determine the association between baseline levels of non-refractory GERD symptoms and new indicators.This study has certain limitations that warrant consideration, particularly the restricted sample size and single-center origin of participants, factors that may compromise the representativeness of the results and their applicability to more diverse demographic populations.

Moreover, a causal relationship between anxiety, depression, sleep disorders and refractory GERD symptoms could not be confirmed.

Overall, a reduction in the distal MNBI to 1531 Ω was found to be helpful in differentiating rGERD from FH and RH. Anxiety, depression, and sleep disturbances have an adverse impact on both the function of the esophageal mucosal barrier and esophageal clearance, which crucially influence rGERD symptoms. Timely evaluation and targeted treatment for anxiety, depression, and sleep disturbances in patients with rGERD thus represents an effective strategy to enhance treatment outcomes.

## Supporting information

S1 Fig.Distal MNBI distinguishing rGERD from FH and RH in the subject operating characteristic curves

S1 Table.Comparison of clinical characteristics of patients in rGERD, FH and RH groups (n=75)

(DOCX)

S2 Table.Comparison of HREM parameters and 24-h impedance-pH monitoring parameters in patients with rGERD, FH, and RH (n=75)

(DOCX)

S3 Table.Comparison of anxiety, depression and sleep quality between patients with rGERD, FH, and RH (n=75)

(DOCX)

S4 Table. Correlations of proximal MNBI, distal MNBI, EGJ-CI, and PSPSW-I with sleep quality, anxiety, and depression

(DOCX)

## Data Availability

The data underlying the results presented in the study are available from Dr.Yang

## Acknowledgements

None.

